# Digital PCR discriminates between SARS-CoV-2 Omicron variants and immune escape mutations

**DOI:** 10.1101/2022.12.19.22283598

**Authors:** Steven C. Holland, LaRinda A. Holland, Matthew F. Smith, Mihyun B. Lee, James C. Hu, Efrem S. Lim

## Abstract

As SARS-CoV-2 continues to evolve, mutations arise that will allow the virus to evade immune defenses and therapeutics. Assays that can identify these mutations can be used to guide personalized patient treatment plans. Digital PCR (dPCR) is a fast and reliable complement to whole genome sequencing that can be used to discriminate single nucleotide polymorphisms (SNPs) in template molecules. Here, we developed a panel of SARS-CoV-2 dPCR assays and demonstrate its applications for typing variant lineages and therapeutic monoclonal antibody resistance. We designed multiplexed dPCR assays for SNPs located at residue 3395 in the *orf1ab* gene and residue 143 of the *spike* gene that differentiate the Delta, Omicron BA.1, and Omicron BA.2 lineages. We demonstrate their validity on 596 clinical saliva specimens that were sequence-verified using Illumina whole genome sequencing. Next, we developed dPCR assays for spike mutations R346T, K444T, N460K, F486V, and F486S mutations that are associated with host immune evasion and reduced therapeutic monoclonal antibody efficacy. We demonstrate that these assays can be run individually or multiplexed to detect the presence of up to 4 SNPs in a single assay. We validate these dPCR assays on 81 clinical saliva SARS-CoV-2 positive specimens from Omicron subvariants BA.2.75.2, BM.1.1, BN.1, BF.7, BQ.1, BQ.1.1, and XBB. Thus, dPCR could serve as a useful tool to determine if clinical specimens contain therapeutically relevant mutations and inform patient treatment.

## INTRODUCTION

The evolution of SARS-CoV-2 brings challenges to disease epidemiology and patient treatment. Genomic mutations arise that define phylogenetic lineages, alter virus properties, and have functional consequences of clinical significance (1). In early November, 2021, the SARS-CoV-2 Delta variant and its sublineages were the predominantly circulating lineages in the United States (2). On November 26, 2021 the World Health Organization designated the SARS-CoV-2 Omicron variant as a “variant of concern” (3). Genome sequencing showed that the Omicron lineages contained approximately 50 unique mutations compared to previous variants of concern (4). Phylogenetic analysis indicated that the Omicron lineages arose independent of the Delta lineage, likely from a prolonged infection in an immunocompromised patient, or an animal host (5). The Omicron BA.1 lineage eventually rose to be the dominant circulating lineage, displacing Delta lineages at a rate faster than previous variants (6). This increased rate of displacement is likely due to increased immune evasion and infectivity of the early Omicron BA.1 and BA.2 variants (7-9).

Since their initial emergence, Omicron lineages have diversified, with multiple sublineages branching from the ancestral BA.1 and BA.2 lineages. Studies show that currently circulating strains (e.g. BA.2.75.2 and BQ.1.1) are increasingly able to escape neutralization by sera obtained from vaccinated individuals, compared to the original BA.2 strain (10). Neutralization by sera from vaccinated individuals that also recovered from an Omicron breakthrough infection (BA.1, BA.2, or BA.5) was also reduced in more recent variants. Worryingly, circulating sublineages have also independently evolved mutations within the receptor binding domain (RBD) of the Spike protein that escape neutralization by the therapeutic monoclonal antibodies (mAbs) bebtelovimab, bamlanivimab, etesevimab, casirivimab, tixagevimab, and others (11). The R346, K444, L452, N460, E484, and F486 residues may be particularly important in mAb resistance and SARS-CoV-2 pathology (12). Due to the limited methods to determine if a patient is infected with a resistant variant, the FDA issues blanket guidance to withdraw emergency use authorization (EUA) for mAbs based on general variant presence (13). This significantly limits patient access to life-saving treatment options regardless of what variant they are infected with (e.g., co-circulating variants that are susceptible to mAbs). Hence, methods to rapidly determine if SARS-CoV-2 positive patient specimens harbor resistance mutations are of critical importance in guiding therapeutic treatment.

Whole genome sequencing is the most comprehensive method for genotyping SARS-CoV-2. However, the cost, time requirements, and required technical expertise leaves a need for complementary methods to identify variants of concern and mutations of interest in clinical specimens. Digital PCR (dPCR) has emerged as a technology that can be used to detect and differentiate single nucleotide polymorphisms (SNPs) in template DNA (14). In the QuantStudio Absolute Q Digital PCR system (ThermoFisher, Massachusetts, USA), template DNA is partitioned into over 20,480 microchambers. Within each microchamber, an end-point PCR reaction is performed which contains primers and probes specific to template DNA. TaqMan (ThermoFisher, Massachusetts, USA) probes contain a fluorescent dye and fluorescent quencher molecule affixed to opposing ends of the probe oligomers. During the PCR elongation phase, polymerase activity degrades annealed probes, liberating the fluorescent dye from the quencher molecule and allowing fluorescence emission (15). Probe length is an important factor in facilitating fluorescence quenching, so minor groove binder (MGB) moieties can also be attached to probes to reduce probe length and improve quencher efficiency (16). After the PCR assay has completed, end-point fluorescence intensity is then used to determine whether a microchamber is ‘positive’ or ‘negative’ for each probe target and the number of positive microchambers can be used to calculate template presence (17). The robustness and sensitivity of dPCR to detect template differences of only a single nucleotide have made it useful in practical applications of viral identification and quantitation, determination of allelic imbalance, and wastewater viral variant surveillance (18-20).

In this study, we demonstrate the usefulness of dPCR in detecting polymorphisms in SARS-CoV-2 genomes obtained from clinical saliva specimens. We show that dPCR can detect the presence of one or two SNPs to distinguish between the Delta, Omicron BA.1, and Omicron BA.2 lineages. We also show that a nine-nucleotide deletion can be used to identify the Omicron BA.1 lineage. We verify the clinical validity of these assays by determining lineage designations for 596 SARS-CoV-2 positive clinical saliva samples and verify those determinations by Illumina next generation sequencing. We finally demonstrate the ability of dPCR to identify mutations at up to 4 genomic loci in a single reaction vessel and use this ability to detect SNPs associated with immune escape in 81 clinical saliva samples.

## MATERIALS AND METHODS

### Saliva specimens and diagnostic testing

This study was approved by Arizona State University Institutional Review Board. This study involved analyses of 677 saliva specimens submitted for SARS-CoV-2 testing at ASU Biodesign Clinical Testing Laboratory (ABCTL) from November 16, 2021 to December 10, 2022. Saliva samples were independently collected by participants in 2 mL collection vials, registered, and deposited at drop-off locations. RNA was extracted from 250µl of saliva specimen within 33 hours of sample receipt using the KingFisher Flex (Thermo Scientific), following the manufacturer’s guidelines. Diagnostic testing was performed using TaqPath COVID-19 Combo Kit assay (Applied Bio-systems, USA) following manufacturer’s guidelines.

### Digital PCR

DNA constructs (primers, probes, template constructs) were ordered from either Integrated DNA technologies (Coralville, IA, USA) or Applied Biosystems (Waltham, MA, USA; nucleotide sequences can be found in Tables S1, S2, and S3 in the supplemental material).

The reaction mixtures for the *orf1ab* and *spike* lineage discriminating assays were comprised of 1X Absolute Q 1-step RT-dPCR Master Mix (Applied Biosystems, Waltham, MA), 400 nM of each primer, 200 nM of each fluorescent probe, 4 μL template, water was used to bring the final volume to 9μL. Synthetic DNAs were assayed at a concentration of 10^3^ copies/μL. Saliva specimens with threshold cycle (C_t_) values less than 19 when assayed using TaqPath COVID-19 Combo Kit assay (Applied Bio-systems, USA) were diluted 1:2 before loading.

The reaction mixtures for the *spike* RBD mutation assays were comprised of 1X Absolute Q 1-step RT-dPCR Master Mix (Applied Biosystems, Waltham, MA), 450 nM of each primer, 280 nM of each fluorescent probe, 5.5 μL template, water was used to bring the final volume to 9μL. Synthetic DNAs were assayed at a concentration of 10^3^ copies/μL.

Samples were loaded in QuantStudio Absolute Q MAP16 (Applied Biosystems, Waltham, MA), overlaid with 15 μL Absolute Q Isolation Buffer (Applied Biosystems, Waltham, MA), and the digital PCR reactions were performed on the Absolute Q dPCR system (Applied Biosystems, Waltham, MA). For the *orf1ab* and *spike* lineage determination assays, cycling conditions were: 10 min at 50°C, 5 min activation at 95°C, 40 cycles of 2s at 95°C and 15s at 54°C. For the *spike* RBD assay determination assays, cycling conditions were: 10 min at 50°C, 5 min activation at 95°C, 40 cycles of 2s at 95°C and 25s at 58°C. Fluorescence intensities and positive chamber counts were analyzed using QuantStudio Absolute Q Digital PCR Software version 6.2.0 (Applied Biosystems, Waltham, MA). Results for clinical specimens can be found in the supplemental material datasets 1 -5.

### SARS-CoV-2 genome sequencing

NGS library preparation for saliva samples was performed using the COVIDSeq Test (Illumina, San Diego, CA, USA) with ARTICv4 and ARTICv4.1 primer sets (21). Libraries were sequenced on the Illumina NextSeq2000 instrument using 2 × 109 paired end reads. Sequencing reads adapter sequences were trimmed using trim-galore, aligned to the Wuhan1 reference genome (MN908947.3) using the Burrows–Wheeler aligner, BWA-MEM version 0.7.17-r1188 (22), and had their primer sequences trimmed using iVAR version 1.3.1 (23). Lineage calling for community and hospital-derived sequencing data was performed with pangolin software (24), with its assignment and designation libraries up to date at the time of analysis. Sequence quality was validated and annotated using VADR version 1.4 (25).

### Phylogenetic analysis

Phylogenies were generated using Nextstrain.cli version 5.0.1 (26) and associated Augur Auspice pipeline with an input dataset based on the Nextstrain ncov GISAID Reference dataset accessed on November 16^th^, 2022. Duplicate lineages were removed, and the set was supplemented with lineages of interest in which assay-targeted mutations were found to be present in at least 75% of sequences. The Nextstrain ncov default build was utilized, with a minimal config file specifying the input dataset and Wuhan/Hu-1/2019 as the root. The filter settings in the default parameters file were modified by setting the skip_diagnostics flag to *true*.

### Global mutation proportion analysis

Global mutation proportion data were downloaded from cov-spectrum (27) with simple queries for S:R346T, S:K444T, S:N460K, S:F486S, and S:F486V. Absolute counts were downloaded in addition to proportion data for S:N460K. An advanced query was performed to obtain absolute counts for each codon (AAA, AAG) present in S:N460K sequences. Data were plotted from March 3^rd^, 2022 until November 6^th^, 2022. For plotted data range, the difference in 95% CI top and 95% CI bottom never reaches greater than 5%)

## RESULTS

### Digital PCR assay to distinguish Delta, Omicron BA.1, and Omicron BA.2 utilizing two SNPs in the *orf1ab* gene

Since the effectiveness of therapeutic treatments can differ between SARS-CoV-2 lineages, a set of dPCR probes was designed to discriminate between the Delta, Omicron BA.1, and BA.2 lineages using SNPs found in the *orf1ab* gene (**Fig 1A**). The probes hybridize at nucleotides 10440-10454, located in the *orf1ab* gene, and are used to detect substitutions found in the Delta and Omicron lineages located at amino acids R3394 and P3395. The probe homologous to the Delta lineage sequence uses VIC as its fluorescent dye and shares the same nucleotide sequence as the Wuhan1 reference sequence over the probe region. The Omicron BA.1 probe discriminates between the Delta sequence at the c10449a nucleotide substitution (amino acid P3395H) and uses FAM as its fluorescent dye. The Omicron BA.2 probe discriminates between the Delta sequence at the c10449a (amino acid P3395H) and g10447a (synonymous mutation in R3394) nucleotide substitutions and uses NED as its fluorescent dye. The fluorescent dyes were connected to the 5’ end of the oligo and all three probes contain a minor groove binder (MGB) moiety on the 3’ end of the oligo to improve binding to template sequence (15).

**Fig 1:**
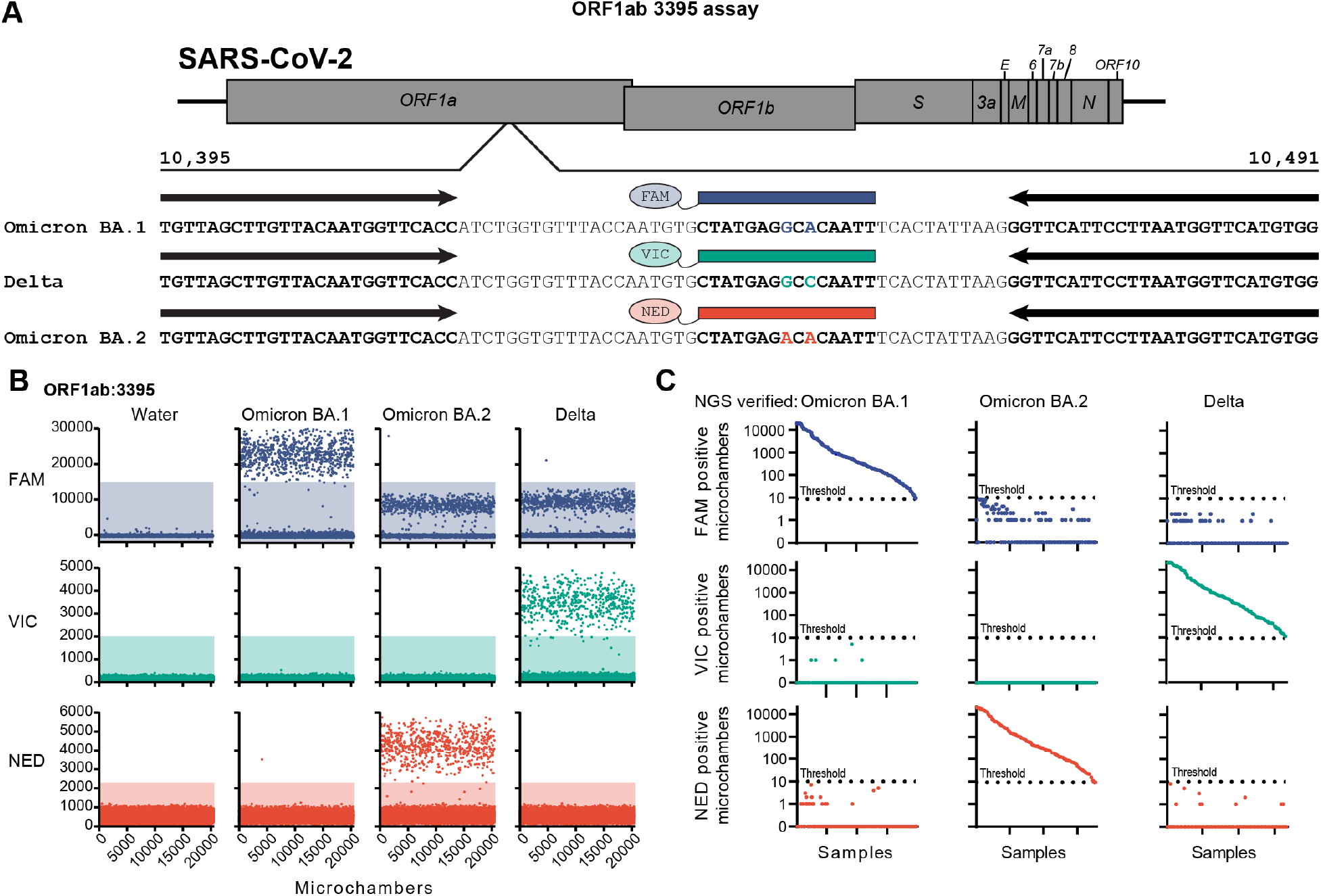
Digital PCR assay for the determination of the Delta, Omicron BA.1, and Omicron BA.2 lineages. (A) Schematic showing annealing locations of primers (black arrows) and probes (colored boxes) on the SARS-CoV-2 genome. (B) Representative fluorescence intensities of dPCR microchambers for synthetic DNA constructs. Positive microchambers are those exceeding fluorescence thresholds (shaded regions). (C) Number of positive microchambers (maximum number of chambers is 20480) resulting from each saliva sample. Samples are grouped by Illumina sequencing lineage determination and sorted by positive microchamber count of the respective lineage-specific probe. Dotted line indicates the positive threshold value (9 microchambers).

The efficacy and specificity of probes were tested using synthetic DNA constructs (**Fig 1B**). The Delta (VIC), Omicron BA.1 (FAM), and Omicron BA.2 (NED) probes showed a clear fluorescence response when assayed with their homologous templates. The Omicron BA.1 probe also showed a fluorescence response to the Delta and Omicron BA.2 templates. This signal was lower in fluorescence intensity than observed from Omicron BA.1 templates, but higher than the negative signal observed in reactions containing no template DNA. There was a negligible response of the Delta (VIC) and Omicron BA.2 (NED) probes when used on their mismatching templates. Fluorescence threshold values for each probe were set to exclude the range of negative values found in mismatching templates. The fluorescence threshold for Omicron BA.1 (FAM) was set above the non-specific fluorescence values observed in the mismatching template assays.

In order to determine the validity of the *orf1ab* dPCR assay on clinical specimens, we performed the 3-probe multiplex dPCR assay and Illumina next-generation sequencing on 596 SARS-CoV-2 positive saliva samples. To receive a ‘positive’ result for a particular probe, the sample required 9 or more microchambers to have a fluorescence intensity above the threshold value. During the dPCR assay scoring, Illumina sample lineages were blinded to scorers. Using this criteria, 540 samples tested positive for a single lineage using the dPCR assay. All 540 samples had lineage designations by dPCR concordant with Illumina whole genome sequencing designations. No samples displayed a positive response from probes discordant to the genome sequencing designations (**Fig 1C;** also see Supplementary Dataset

1 in the supplemental material). There were 56 samples in which no probe passed the positive chamber count. C_t_ values of these negative samples were found to be significantly higher than samples with positive outcomes, indicating that viral load contributes to assay efficacy (Positive samples median C_t_ = 22.05, Negative samples median C_t_ = 24.34, *p* < 0.0001; Mann-Whitney test). Together, this demonstrates that dPCR can use two nucleotide polymorphisms to discriminate the Delta, Omicron BA.1, and Omicron BA.2 lineages in saliva specimens.

### Digital PCR assay to distinguish between Omicron BA.1 and Delta/Omicron BA.2 utilizing a nine-nucleotide deletion in the *spike* gene

Some therapeutic antibodies, like sotrovimab, are still partially effective against the Omicron BA.1 lineage but less effective against the BA.2 lineage (11), we sought to design an assay to discriminate the Omicron BA.1 lineage from Delta and BA.2. We found a nine-nucleotide deletion within the Spike protein coding region (G21987-T21995, amino acids G142-Y145) specific to the BA.1 lineage. We designed dPCR probes to either contain only the nucleotides flanking the deleted region or to contain the nucleotides found in the deletion (**Fig 2A**). These sequences would provide identity to the Omicron BA.1 lineage, or to the Delta and Omicron BA.2 lineages, respectively. Probes contained either the VIC (BA.1) or FAM (Delta/BA.2) fluorescent dye on the 5’ end and both probes contained a 3’ MGB moiety.

**Fig 2:**
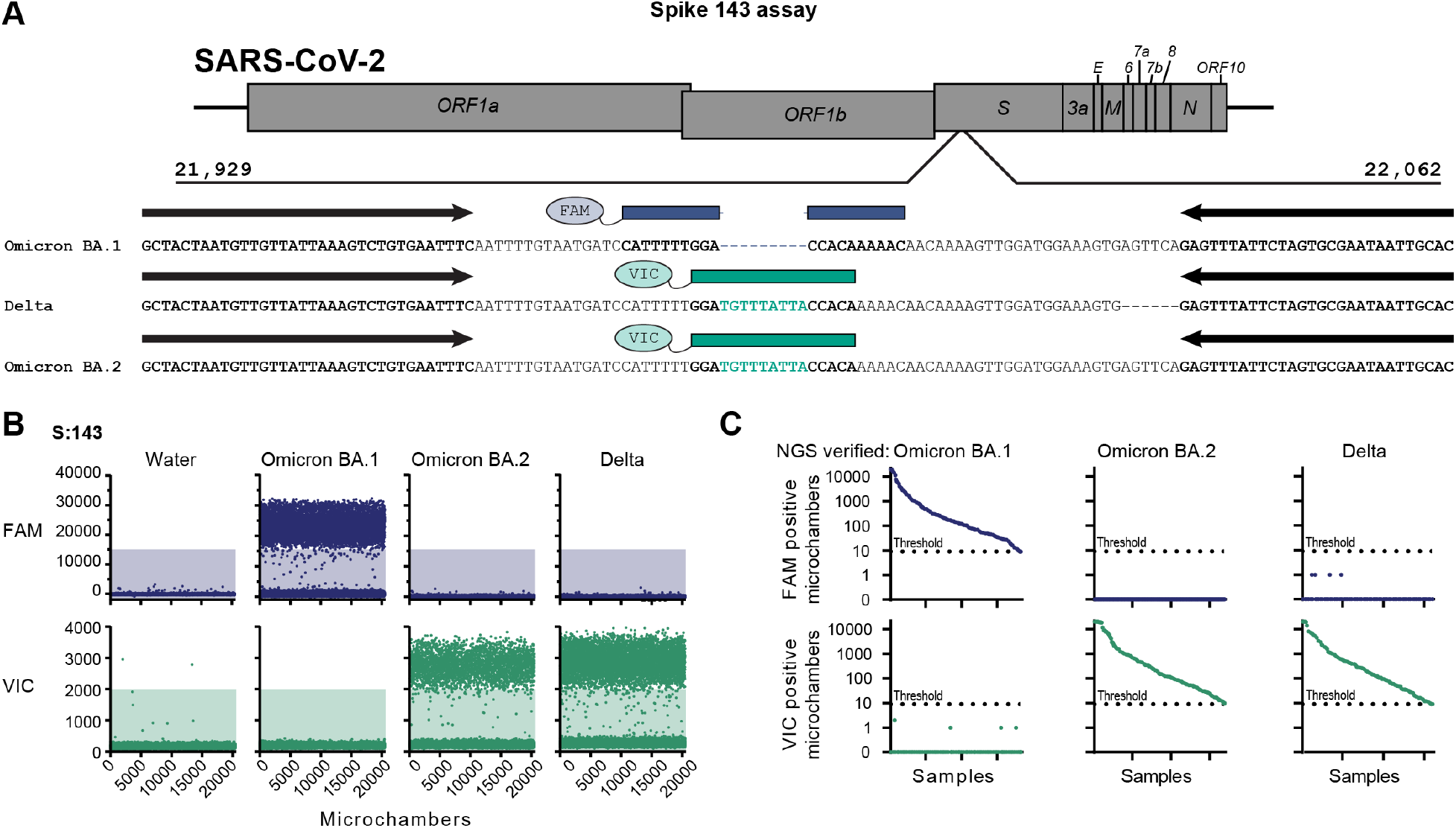
Digital PCR assay for the determination of the Omicron BA.1, and Delta or Omicron BA.2 lineages. (A) Schematic showing annealing locations of primers (black arrows) and probes (colored boxes) on the SARS-CoV-2 genome. (B) Representative fluorescence intensities of dPCR microchambers for synthetic DNA constructs. Positive microchambers are those exceeding fluorescence thresholds (shaded regions). (C) Number of positive microchambers (maximum number of chambers is 20480) resulting from each saliva sample. Samples are grouped by Illumina sequencing lineage determination and sorted by positive microchamber count of the respective lineage-specific probe. Dotted line indicates the positive threshold value (9 microchambers).

The dPCR probes were validated on synthetically produced DNA templates. Each probe displayed a high fluorescence signal when assayed with its matching template (**Fig 2B**). No evidence of a secondary fluorescence signal band, as seen in the *orf1ab* Omicron BA.1 probe (**Fig 1B**), was observed in either probe (**Fig 2B**). Fluorescence thresholds were set by excluding fluorescence intensity values from mismatching control templates.

The S gene dPCR assay was performed on the same 596 SARS-CoV-2 positive clinical saliva samples assayed on the *orf1ab* assay. Sample positivity was scored as above, blinded and using the 9 microchambers as the positive threshold criterion. Using this method, 512 samples tested positive from a single probe. All 512 lineage designations from Illumina whole genome sequencing were in concordance with the digital PCR assay. No sample tested positive to a dPCR probe that was discordant to its lineage designation from whole genome sequencing (**Fig 2C;** also see Supplementary Dataset 2 in the supplemental material). There were 84 samples in which no probe passed the positive threshold value. C_t_ values of samples with no positive results were found to be significantly higher than positive samples, indicating that viral load contributes to assay efficacy (Positive samples median C_t_ = 21.88, Negative samples median C_t_ = 24.42, *p* < 0.0001; Mann-Whitney test). This demonstrates that the S gene del143-145 deletion can be used to discriminate SARS-CoV-2 Omicron BA.1 lineages from Delta and Omicron BA.2 lineages in saliva specimens.

### Digital PCR assays to detect immune evasion-associated SNPs in the Spike protein receptor binding domain

Another potential diagnostic application is in discerning the presence of amino acid mutations that may render therapeutic monoclonal antibodies ineffective. To observe the frequency of Spike RBD mutations that provide immune escape properties, the GISAID database was queried for SARS-CoV-2 Omicron sequences containing five mutations associated with immune escape: R346T, K444T, N460K, F486V, and F486S (10) (**Fig 3**). The L452R mutation within the Spike RBD region has also been strongly associated with immune escape (10), but has been the subject RT-qPCR detection (28), so it was omitted from analysis. We found that many of these mutations are found distributed across multiple sublineages (**Fig 3A**). We observed that the R346T mutation has been rising in frequency since May 2022 and has arisen in the BA.2, BA.4, and BA.5 lineages as well as in the XBB recombinant lineages (**Fig 3B)**. The K444T mutation has been rising in global frequency since September 2022, arising within the BE and BQ sublineages (BE and BQ are sublineages of BA.5; **Fig 3C**). The N460K mutation has arisen within BA.2 and BA.5 sublineages and XBB recombinants (**Fig 3D**). The BA.2 sublineages utilize an AAG codon, whereas the BA.5 lineages utilize an AAA codon. The F486S mutation is slowly becoming more abundant and is seen in BA.2 sublineages and XBB recombinants (**Fig 3E**). The F486V mutation arose earliest and differentiates BA.4 and BA.5 lineages from BA.2 lineages (**Fig 3F**). Taken together, these mutations are becoming more abundant in circulating SARS-CoV-2 lineages and are arising in multiple, independent lineages.

**Fig 3:**
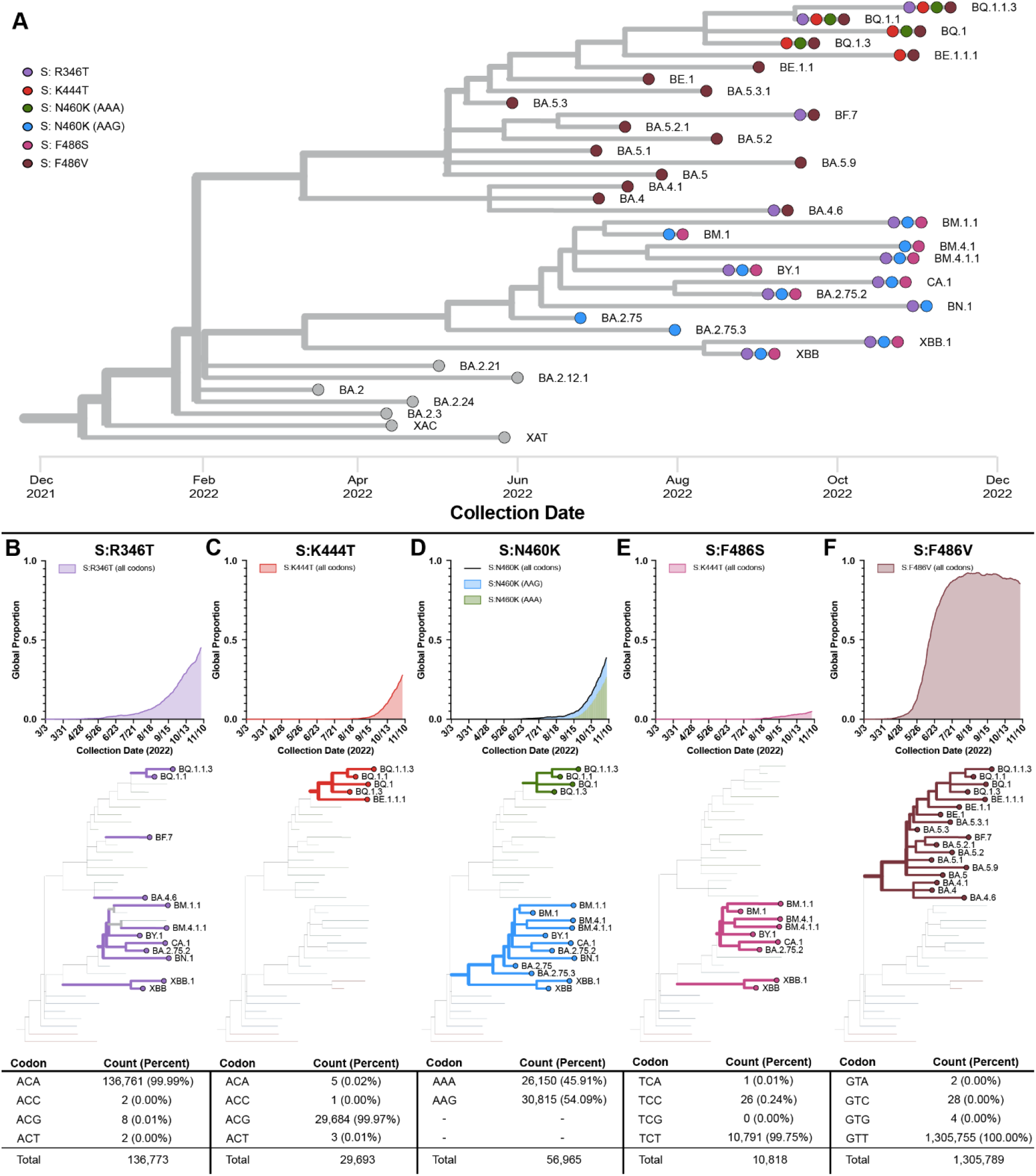
Phylogenetic distribution of mutations that evade therapeutic monoclonal antibodies. (A) Phylogenetic tree of representative circulating sublineages. Colored circles indicate the presence of a Spike RBD mutation found in at least 75% of sequences within that lineage. Global frequency, phylogenetic distribution, and codon frequencies for R346T (B), K444T (C), N460K (D), F486S (E), and F486V (F) mutations.

To ensure that dPCR probes were designed with sequences representative of circulating SARS-CoV-2 genomes, we queried the GISAID database to determine the codon usage for amino acid mutations associated with immune escape. For the R346T, K444T, F486V and F486S mutations, over 99% of genome sequences in the GISAID database utilize a single codon to encode the substituted amino acid (**Figs 3A, 3B, 3C, 3E, 3F**). However, N460K is encoded by the AAG codon in 54.09% of sequences and by the AAA codon in 45.91% of genomes containing this mutation (**Fig 3D**). Therefore, two probes were designed to investigate the N460K mutation. Probes containing the amino acid found in the Wuhan1 reference sequence at each position were also created at each position. To assess the global applicability of the dPCR assays, we calculated the mismatch frequency at each nucleotide position for all primers and probes against all genome sequences in the GISAID database. We found that probes had low mismatch frequency outside of the codon encoding the mutation of interest (see Fig S1 in the supplemental material). Primers also had a low mismatch frequency, however, the F486 assay requires the presence of the S477N and T478K mutations for amplicon generation.

Mutation and reference probes were designed to be run independently or multiplexed with other probes. Each mutation/reference pair contained a different fluorescent dye and the mutation probes used a different fluorescent dye between residue locations (**Fig 4A**). Mutation probes for sites containing two mutations (i.e. N460K (AAA) and N460K (AAG), F486S and F486V) use the same fluorescent dye for each mutation. The R346 and F486 probe sets use unique, independent amplicons, whereas the K444 and N460 probe sets share a single, common amplicon.

**Fig 4:**
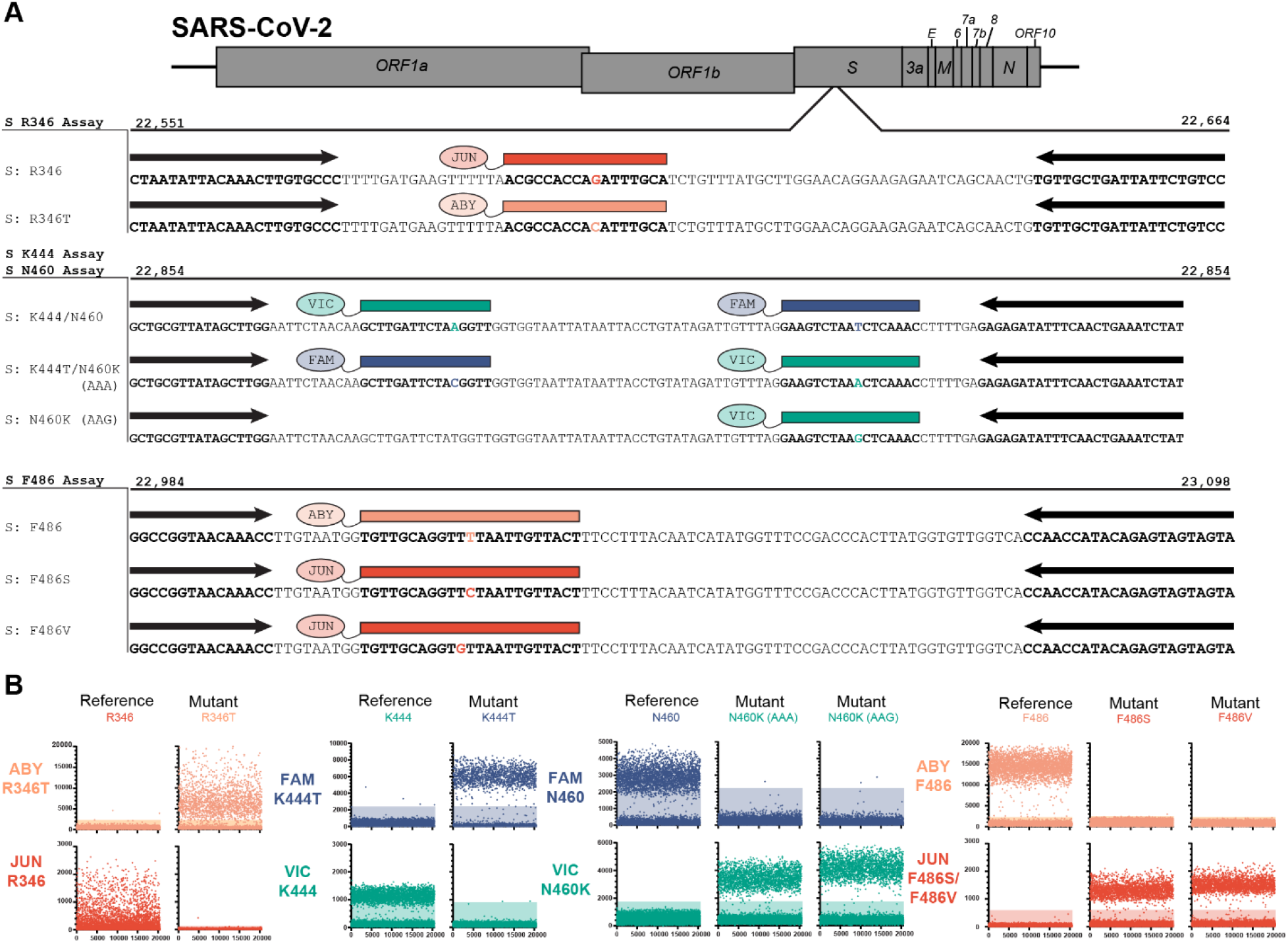
Digital PCR assay for the detection of mutations associated with therapeutic monoclonal antibody evasion. (A) Schematic showing annealing locations of primers (black arrows) and probes (colored boxes) on the SARS-CoV-2 genome. (B) Representative fluorescence intensities of dPCR microchambers for synthetic DNA constructs. Positive microchambers are those exceeding fluorescence thresholds (shaded regions).

In order to determine the effectiveness of the dPCR probes to discriminate the identity of each SNP, reactions containing the probe homologous to the mutation of interest and a probe homologous to the reference sequence were assayed in a single reaction vessel. Each probe pair was assayed using synthetically produced DNA templates containing the mutation nucleotide sequence or reference nucleotide sequence. For each reaction, each probe displayed a higher fluorescence signal to its homologous template, than to a template containing a mismatching nucleotide (**Fig 4B**). Positive FAM fluorescence was observed from assays with templates containing N460 or K444T residues. Positive VIC fluorescence was observed from assays with templates containing N460K or K444 residues. Positive ABY fluorescence was observed from assays with templates containing R346T or F486 residues. Positive JUN fluorescence was observed from assays using templates containing R346, F486S, or F486V templates. Together, this demonstrates that each probe can correctly discriminate templates differing by only a single SNP within the probe region.

The primers and probes from two assays were combined to assay the identity of two SNPs from a single template. For each reaction, two amplicons were produced, with each amplicon containing only a single substitution of interest. When assayed on synthetic DNA constructs, each probe in the assays displayed higher fluorescence to its homologous template, than to a mismatching template. The R346 and K444 assays were combined into single dPCR assay to interrogate sequence identity at those positions simultaneously (**Fig 5A**). On the template containing reference sequences, positive fluorescence was observed in VIC and JUN channels. A positive response was observed in FAM and ABY channels for a template containing K444T and R346T mutations, respectively.

**Fig 5:**
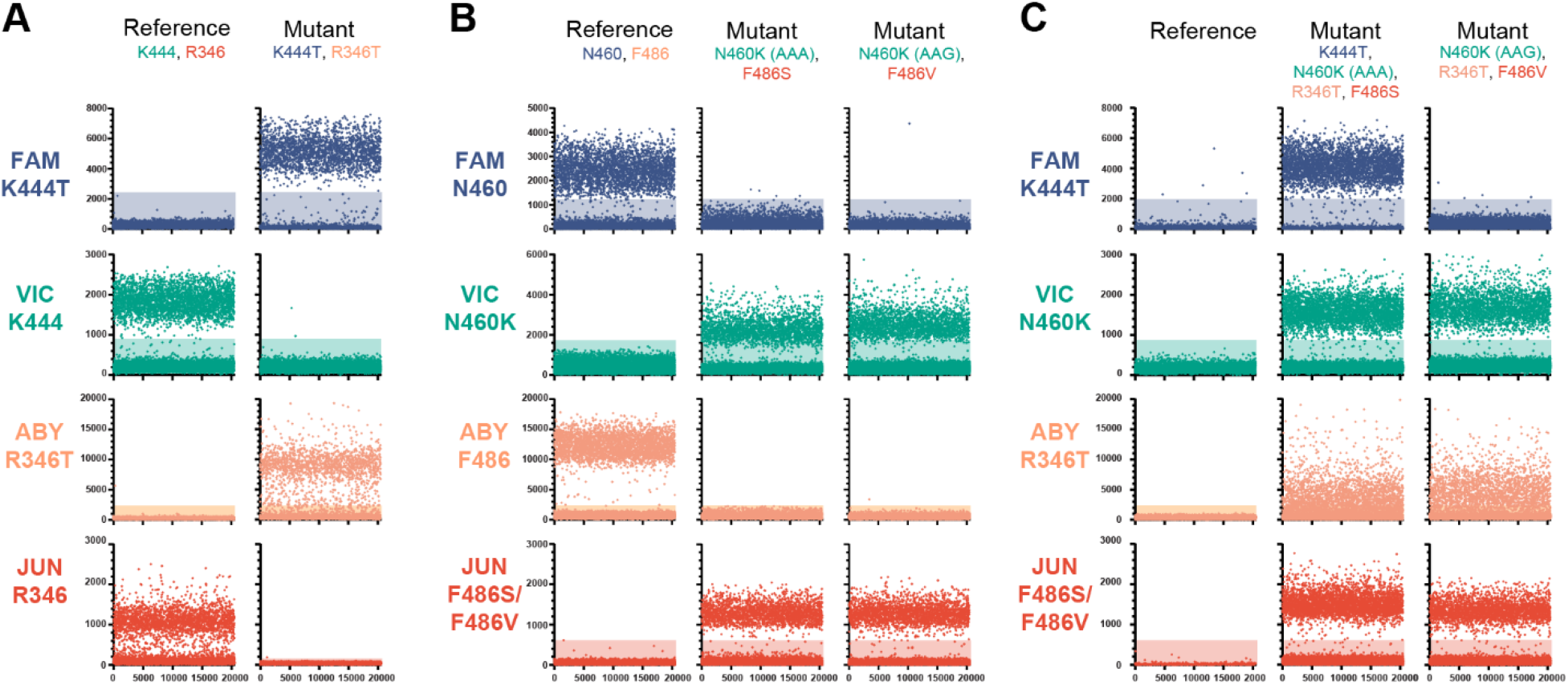
Representative microchamber fluorescence intensities for synthetic DNA constructs from multiplexed dPCR assays. (A) R346 and K444 combination assay. (B) N460 and F486 combination assay. (C) Combination assay using R346T, K444T, N460K (AAA and AAG codons), F486S, and F486V mutation probes. Positive microchambers are those exceeding fluorescence thresholds (shaded regions).

The N460 and F486 assays were also combined into a single assay (**Fig 5B**). The reaction contained a labeled probe for each reference sequence, a VIC-labeled probe for each N460K codon (AAG and AAA), and a JUN-labeled probe for each F486 mutation (F486S and F486V). For the reference sequence template, positive fluorescence responses were observed in the FAM and ABY channels, and for both mutation templates positive responses were observed in the VIC and JUN channels. An equivalent positive response was observed for the N460K AAG and AAA codon probes as well as between the F486S and F486V probes, for their respective templates.

The presence or absence of mutations at 4 SNP sites was able to be resolved in a single dPCR reaction by using the mutation specific probes containing a different fluorescent dye for each locus. Three amplicons were produced in each reaction, with the K444 and N460 probes sharing an amplicon. For each probe, fluorescence values were higher for its homologous template than to its mismatching template (**Fig 5C**). No positive response from any channel was observed to a template containing the reference sequence at each position. Positive responses in the VIC, ABY, and JUN channels were observed on a template containing N460K (AAG), R346T, and F486V mutations. Positive responses in all four channels were observed on a template containing the K444T, N460K (AAA), R346T, and F486S mutations. The number of positive microchambers were similar between templates and probes, excepting the R346T probe having a lower positive chamber count. These results demonstrate the versatility of dPCR assays to report the identity of multiple nucleotide sequences across a template.

In order to test the validity of these multiplexed RT-dPCR assays on clinical samples, we performed dPCR and Illumina sequencing assays on human saliva samples. Saliva samples containing SARS-CoV-2 infections from 8 lineages, previously identified by Illumina whole genome sequencing, were used to evaluate the specificity of the assay to discriminate among clinical specimens with varying mutation profiles. For each lineage, samples were examined using an assay where the R346 and K444 assays were combined, an assay where the N460 and F486 assays were combined, and an assay where all 6 mutation-specific probes were used. The combined R346 and K444 assay was able to correctly discriminate between the mutation and reference sequence at each locus (**Fig 6A**). FAM fluorescence was observed in the BQ.1 and BQ.1.1 lineages, which contain the K444T mutation. Positive VIC fluorescence was seen in samples from the BA.2, BA.2.75.2, BM.1.1, BN.1, BA.5, and BF.7 lineages, consistent with the presence of the reference sequence at the K444 locus. While the XBB lineage contains the reference amino acid at K444 and V445P mutation. The V445P mutation is not present in the reference-specific probe and causes failure at this position. A positive ABY response was seen in the BA.2.75.2, BM.1.1, BN.1, BF.7, BQ.1.1, and XBB lineages, which contain the R346 mutation. Finally, JUN fluorescence properly identified the presence of the reference sequence at R346 in the BA.2, BA.5, and BQ.1 lineages. A reduced positivity response of the R346 probes was also observed on the saliva samples, as seen with the synthetic DNA constructs. Thirty-two total samples were tested on the combined assay and gave results consistent with their Illumina whole genome sequencing determination (see Supplementary Dataset 3 in the supplemental material).

**Fig 6:**
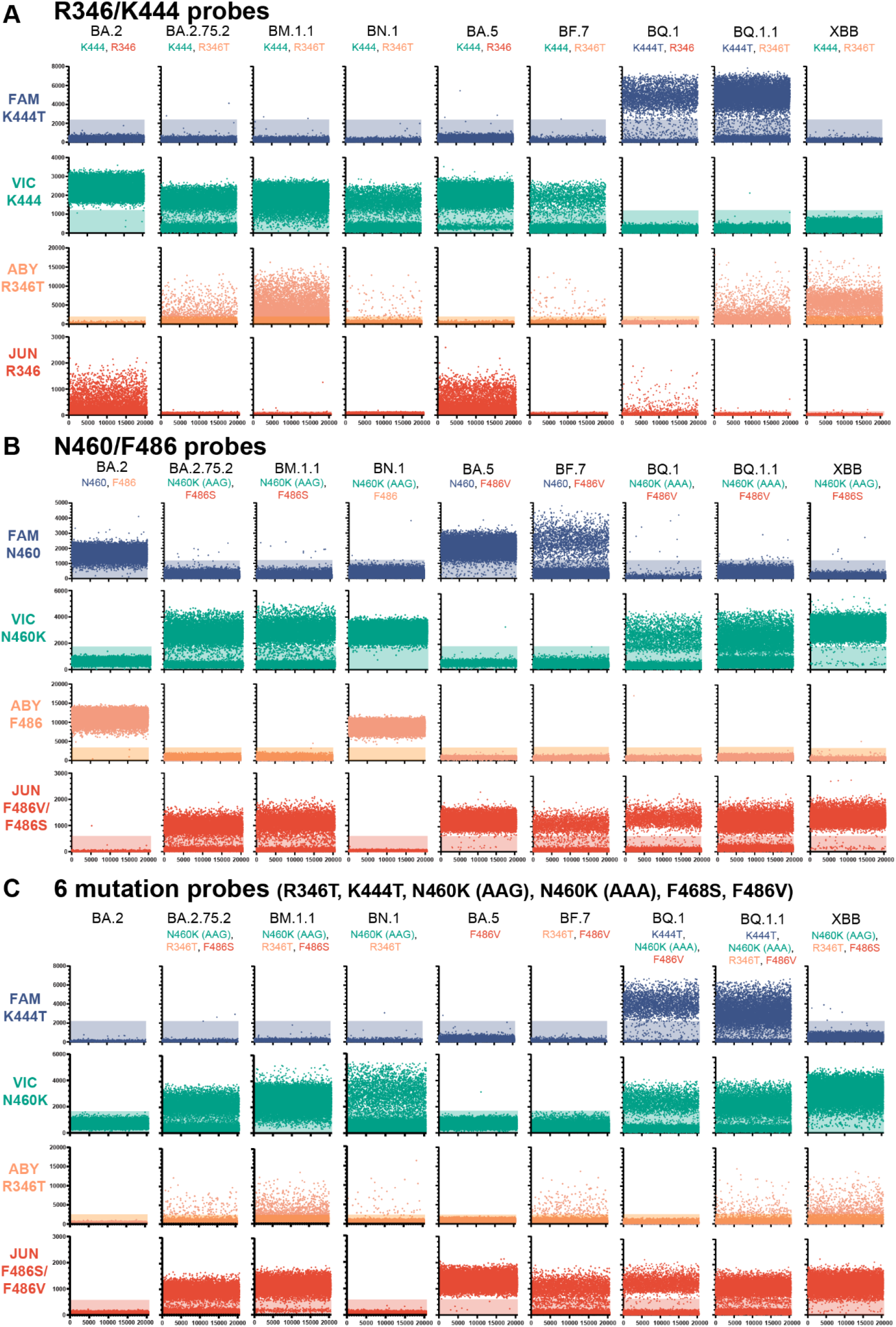
Representative microchamber fluorescence intensities for clinical saliva specimens from multiplexed dPCR assays. (A) R346 and K444 combination assay. (B) N460 and F486 combination assay. (C) Combination assay using R346T, K444T, N460K (AAA and AAG codons), F486S, and F486V mutation probes. Positive microchambers are those exceeding fluorescence thresholds (shaded regions).

Sequence composition of saliva samples was correctly identified using the N460 and F486 combined assays (**Fig 6B**). Positive FAM fluorescence was observed in the BA.2, BA.5, and BF.7 lineages, which contain the reference nucleotide sequence at N460. Positive VIC fluorescence was observed in the BA.2.75.2, BM.1.1, BN.1, BQ.1, BQ.1.1, and XBB lineages, consistent with the presence of the N460K mutation in these lineages. Positive ABY fluorescence was observed in the BA.2 and BA.5 lineages, which have the reference nucleotide sequence at F486. Positive JUN fluorescence was observed in BA.2.75.2, BM.1.1, BA.5, BF.7, BQ.1, BQ.1.1, and XBB lineages, which contain either the F486S or F486V mutations. Twenty-eight total samples were tested on the combined assay and gave results consistent with their Illumina whole genome sequencing determination (see Supplementary Dataset 4 in the supplemental material).

An assay comprised of the 6 mutation-specific probes was also verified on saliva samples and gave appropriate responses to mutation composition (**Fig 6C**). Positive FAM fluorescence was observed in the BQ.1 and BQ.1.1 lineages, consistent with the presence of the K444T mutation. Positive VIC fluorescence was observed in the BA.2.75.2, BM.1.1, BN.1, BQ.1, BQ.1.1, and XBB lineages, consistent with the presence of either N460K mutation (AAG or AAA codon). Positive ABY fluorescence was observed in the BA.2.75.2, BM.1.1, BN.1, BF.7, BQ.1.1, and XBB lineages, consistent with the presence of the R346T mutation. Positive JUN fluorescence was observed in the BA.2.75.2, BM.1.1, BA.5, BF.7, BQ.1, BQ.1.1, and XBB lineages, consistent with the presence of either the F486S or F486V mutations. No positive fluorescence response was observed in the BA.2 lineage, which contains none of the screened mutations. As in the R346/K444 combination assay, the XBB lineage failed to display FAM fluorescence due to the absence of the K444T mutation and the presence of the V445P mutation. We tested this assay on 75 total saliva samples, and all samples displayed results consistent with their respective Illumina whole genome sequencing lineage designation (see Supplementary Dataset 5 in the supplemental material). These assays illustrate that dPCR can discern immunologically relevant SARS-CoV-2 mutations from saliva specimens and could be used to guide therapeutic treatment.

## DISCUSSION

In this study we demonstrated the robustness of using digital PCR in genotyping SARS-CoV-2 positive saliva samples for two use cases: lineage classification and therapeutic mAb resistance. We showed that using a mixture of probes with a common fluorescent dye can be used to identify mutants when multiple nucleotide substitutions are at one region of interest (N460 and F486 assays, **Fig 4B**). This is particularly useful when an amino acid is encoded by multiple codons, or multiple amino acids are of interest. We also demonstrated that detection is not compromised when multiple probes bind to different locations on a single amplicon (N460/F486 assay, **Fig 5C**). This property is beneficial when interrogating multiple mutations within a single domain of a protein (e.g. the RBD of the *s* gene) where creating multiple amplicons could be difficult due to primer constraints. Finally, we demonstrated that these assays can be performed on nucleic acids extracted from saliva samples to detect the presence of specific SARS-CoV-2 mutations.

The R346, K444, N460, and F486 residues are locations of key mutations important in neutralization escape, enhanced fusogenicity, and enhanced S protein processing. Structural modeling suggests that R346T and K444T appear to disrupt salt bridge formation between S protein and class III monoclonal antibodies (e.g. cilgavimab), lowering effectiveness (12, 29). The N460K mutation has been shown to enhance S protein processing and is predicted to facilitate a salt bridge and hydrogen bond formation between Spike and human ACE2, enhancing cell fusion (30). The F486V mutation aided in evading serum neutralization by early BA.5 lineages, but more recent lineages containing the F486S mutation are even more resistant to neutralization (31). Beyond the mutations examined here, additional mutations at these loci have been implicated in immune escape and are rising in frequency (11).

There are limitations to the use of dPCR for SNP identification. Since dPCR relies on sequence homology between probes, primers, and template nucleic acids, it is susceptible to failure as SARS-CoV-2 evolves mutations. There have been numerous reports of SARS-CoV-2 mutations that cause failure on diagnostic RT-PCR assays (32-34). Mutations in other nucleotides of the probe or primer binding regions will result in failed amplification, as observed with the K444 reference probe against the XBB lineage (**Fig 6A**), and could result in sequence misclassification. Furthermore, caution must be exercised interpreting negative results of reactions containing only a single probe for each SNP of interest (i.e. 6 mutant probe, 4 SNP assay; **Fig 6C**). Results from lineages containing no mutations of interest, such as BA.2, are indistinguishable from failed assays and samples with concentrations below the limit of detection. This limitation could be overcome using a universally binding probe that would always provide a fluorescent response but doing so would reduce the total number of SNP sites being interrogated in a single reaction.

As new variants emerge, the effectiveness of current antibody therapies is at risk. At their emergence, the initial Omicron variants were less sensitive to antibody treatments bamlanivimab and etesevimab (joint administration) and REGEN-COV (casirivimab and imdevimab), and their use was curtailed (35). More recent variants have caused the recall of emergency use authorization for bebtelovimab, leaving Paxlovid (nirmatrelivir/ritonavir) and remdesivir as preferred treatment options (36). These treatments are also at risk, as *in vitro* analysis using a Vesicular Stomatitis Virus system has identified that Y54C, G138S, L167F, Q192R, A194S and F305L mutations in nsp5 (3-cymotrypsin like protease) could result in reduced Paxlovid efficacy (37). As demonstrated in this study, a similar dPCR approach could be used to determine resistance to Paxlovid. Due to the very rapid development of resistance mutations in SARS-CoV-2 (38) there is an increased need for versatile assays to guide patient treatment and extend the use of life-saving therapeutics. The use of digital PCR to clarify active infection (39), determine SARS-CoV-2 lineages, and pinpoint therapeutically relevant mutations make it a well-suited technology for use in personalized diagnostics.

## Data Availability

Genome sequences have been deposited to the GISAID repository. Scripts for data curation can be found on the Lim lab GitHub (https://github.com/ASU-Lim-Lab/Absolute-Q).

## ACKNOWLEDGEMENTS

We thank the authors from originating laboratories responsible for obtaining the specimens and the submitting laboratories where genetic sequence data were generated and shared via the GISAID initiative, on which part of the research is based.

## ABBREVIATIONS

(SARS-CoV-2): Severe acute respiratory syndrome coronavirus 2
(COVID-19): Coronavirus Disease 2019
(RT-dPCR): Reverse transcription-digital polymerase chain reaction
(VOC): Variant of concern

## AUTHOR CONTRIBUTIONS

Conceptualization: S.C.H. and E.S.L.; data curation: S.C.H., L.A.H., M.B.L., J.C.H, M.F.S; formal analysis: S.C.H., L.A.H., M.F.S.; funding acquisition: E.S.L.; investigation: S.C.H., L.A.H., M.L., M.F.S., J.C.H.; supervision: E.S.L.; writing – original draft: S.C.H. and E.S.L.; writing – review and editing S.C.H. and E.S.L. All authors have read and agreed to the published version of the manuscript.

## FIGURE LEGENDS

**Fig S1**: Failure frequency of probes (A) and primers (B) used in the *spike* RBD digital PCR assays.

**Table S1**: Digital PCR probe sequences and modifications

**Table S2**: Digital PCR primer sequences

**Table S3**: Digital PCR synthetic DNA templates used in

**Supplementary Dataset 1**: Number of positive wells passing threshold value and metadata for each specimen in the Orf1ab_3395 assay

**Supplementary Dataset 2**: Number of positive wells passing threshold value and metadata for each specimen in the spike 143 deletion assay

**Supplementary Dataset 3**: Number of positive wells passing threshold value and metadata for each specimen in the R346/K444 combined assay

**Supplementary Dataset 4**: Number of positive wells passing threshold value and metadata for each specimen in the N460/F486 combined assay

**Supplementary Dataset 5**: Number of positive wells passing threshold value and metadata for each specimen in the 6 mutation probe assay

## REFERENCES

1. Tao K, Tzou PL, Nouhin J, Gupta RK, de Oliveira T, Kosakovsky Pond SL, Fera D, Shafer RW. 2021. The biological and clinical significance of emerging SARS-CoV-2 variants. Nat Rev Genet 22:757–773.

2. Centers for Disease Control and Prevention. 2022. COVID Data Tracker, on US Department of Health and Human Services, DC. https://covid.cdc.gov/covid-data-tracker. Accessed 2022-12-13.

3. World Health Organization. 2021. Enhancing Readiness for Omicron (B.1.1.529): Technical Brief and Priority Actions for Member States. https://www.who.int/docs/default-source/coronaviruse/technical-brief-and-priority-action-on-omicron.pdf?sfvrsn=50732953_3. Accessed 2022/11/14.

4. Du P, Gao GF, Wang Q. 2022. The mysterious origins of the Omicron variant of SARS-CoV-2. Innovation (Camb) 3:100206.

5. Sun Y, Lin W, Dong W, Xu J. 2022. Origin and evolutionary analysis of the SARS-CoV-2 Omicron variant. J Biosaf Biosecur 4:33–37.

6. Smith MF, Holland SC, Lee MB, Hu JC, Pham NC, Sullins RA, Holland LA, Mu T, Thomas AW, Fitch R, Driver EM, Halden RU, Villegas-Gold M, Sanders S, Krauss JL, Nordstrom L, Mulrow M, White M, Murugan V, Lim ES. in press. Baseline sequencing surveillance of public clinical testing, hospitals, and community wastewater reveals rapid emergence of SARS-CoV-2 Omicron variant of concern in Arizona, USA. mBio.

7. Zhang X, Wu S, Wu B, Yang Q, Chen A, Li Y, Zhang Y, Pan T, Zhang H, He X. 2021. SARS-CoV-2 Omicron strain exhibits potent capabilities for immune evasion and viral entrance. Signal Transduct Target Ther 6:430.

8. McCallum M, Czudnochowski N, Rosen LE, Zepeda SK, Bowen JE, Walls AC, Hauser K, Joshi A, Stewart C, Dillen JR, Powell AE, Croll TI, Nix J, Virgin HW, Corti D, Snell G, Veesler D. 2021. Structural basis of SARS-CoV-2 Omicron immune evasion and receptor engagement. Science 375:864–868.

9. Chen J, Wang R, Gilby NB, Wei GW. 2022. Omicron Variant (B.1.1.529): Infectivity, Vaccine Breakthrough, and Antibody Resistance. J Chem Inf Model 62:412–422.

10. Cao Y, Jian F, Wang J, Yu Y, Song W, Yisimayi A, Wang J, An R, Chen X, Zhang N, Wang Y, Wang P, Zhao L, Sun H, Yu L, Yang S, Niu X, Xiao T, Gu Q, Shao F, Hao X, Xu Y, Jin R, Shen Z, Wang Y, Xie XS. 2022. Imprinted SARS-CoV-2 humoral immunity induces converging Omicron RBD evolution. bioRxiv doi:10.1101/2022.09.15.507787.

11. Focosi D, McConnell S, Casadevall A. 2022. The Omicron variant of concern: Diversification and convergent evolution in spike protein, and escape from anti-Spike monoclonal antibodies. Drug Resist Updat 65:100882.

12. Qu P, Evans JP, Faraone JN, Zheng YM, Carlin C, Anghelina M, Stevens P, Fernandez S, Jones D, Lozanski G, Panchal A, Saif LJ, Oltz EM, Xu K, Gumina RJ, Liu SL. 2022. Enhanced neutralization resistance of SARS-CoV-2 Omicron subvariants BQ.1, BQ.1.1, BA.4.6, BF.7, and BA.2.75.2. Cell Host Microbe doi:10.1016/j.chom.2022.11.012.

13. US FDA. 2022. FDA announces bebtelovimab is not currently authorized in any US region. https://www.fda.gov/drugs/drug-safety-and-availability/fda-announces-bebtelovimab-not-currently-authorized-any-us-region. Accessed 2022-12-11.

14. Butchbach ME. 2016. Applicability of digital PCR to the investigation of pediatric-onset genetic disorders. Biomol Detect Quantif 10:9–14.

15. Navarro E, Serrano-Heras G, Castano MJ, Solera J. 2015. Real-time PCR detection chemistry. Clin Chim Acta 439:231–50.

16. Kutyavin IV, Afonina IA, Mills A, Gorn VV, Lukhtanov EA, Belousov ES, Singer MJ, Walburger DK, Lokhov SG, Gall AA, Dempcy R, Reed MW, Meyer RB, Hedgpeth J. 2000. 3’-minor groove binder-DNA probes increase sequence specificity at PCR extension temperatures. Nucleic Acids Res 28:655–61.

17. Jiang L, Lin R, Gallagher S, Zayac A, Butchbach MER, Hung P. 2020. Development and validation of a 4-color multiplexing spinal muscular atrophy (SMA) genotyping assay on a novel integrated digital PCR instrument. Sci Rep 10:19892.

18. Sedlak RH, Cook L, Cheng A, Magaret A, Jerome KR. 2014. Clinical utility of droplet digital PCR for human cytomegalovirus. J Clin Microbiol 52:2844–8.

19. Lo YM, Lun FM, Chan KC, Tsui NB, Chong KC, Lau TK, Leung TY, Zee BC, Cantor CR, Chiu RW. 2007. Digital PCR for the molecular detection of fetal chromosomal aneuploidy. Proc Natl Acad Sci U S A 104:13116–21.

20. Caduff L, Dreifuss D, Schindler T, Devaux AJ, Ganesanandamoorthy P, Kull A, Stachler E, Fernandez-Cassi X, Beerenwinkel N, Kohn T, Ort C, Julian TR. 2022. Inferring transmission fitness advantage of SARS-CoV-2 variants of concern from wastewater samples using digital PCR, Switzerland, December 2020 through March 2021. Euro Surveill 27.

21. ARTIC Network. 2022. artic-network/primer-schemes/nCOV-2019. https://github.com/artic-network/primer-schemes/tree/master/nCoV-2019/V4. Accessed 2022-12-01.

22. Li H. 2013. Aligning sequence reads, clone sequences and assembly contigs with BWA-MEM. arXiv 1303.3997.

23. Castellano S, Cestari F, Faglioni G, Tenedini E, Marino M, Artuso L, Manfredini R, Luppi M, Trenti T, Tagliafico E. 2021. iVar, an Interpretation-Oriented Tool to Manage the Update and Revision of Variant Annotation and Classification. Genes (Basel) 12.

24. Rambaut A, Holmes EC, O’Toole A, Hill V, McCrone JT, Ruis C, du Plessis L, Pybus OG. 2020. A dynamic nomenclature proposal for SARS-CoV-2 lineages to assist genomic epidemiology. Nat Microbiol 5:1403–1407.

25. Schaffer AA, Hatcher EL, Yankie L, Shonkwiler L, Brister JR, Karsch-Mizrachi I, Nawrocki EP. 2020. VADR: validation and annotation of virus sequence submissions to GenBank. BMC Bioinformatics 21:211.

26. Hadfield J, Megill C, Bell SM, Huddleston J, Potter B, Callender C, Sagulenko P, Bedford T, Neher RA. 2018. Nextstrain: real-time tracking of pathogen evolution. Bioinformatics 34:4121–4123.

27. Chen C, Nadeau S, Yared M, Voinov P, Xie N, Roemer C, Stadler T. 2021. CoV-Spectrum: Analysis of Globally Shared SARS-CoV-2 Data to Identify and Characterize New Variants. Bioinformatics 38:1735–7.

28. Jessen R, Nielsen L, Larsen NB, Cohen AS, Gunalan V, Marving E, Alfaro-Nunez A, Polacek C, Danish C-GC, Fomsgaard A, Spiess K. 2022. A RT-qPCR system using a degenerate probe for specific identification and differentiation of SARS-CoV-2 Omicron (B.1.1.529) variants of concern. PLoS One 17:e0274889.

29. Wang Q, Li Z, Ho J, Guo Y, Yeh AY, Mohri H, Liu M, Wang M, Yu J, Shah JG, Chang JY, Herbas F, Yin MT, Sobieszczyk ME, Sheng Z, Liu L, Ho DD. 2022. Resistance of SARS-CoV-2 omicron subvariant BA.4.6 to antibody neutralisation. Lancet Infect Dis 22:1666–1668.

30. Qu P, Evans JP, Zheng YM, Carlin C, Saif LJ, Oltz EM, Xu K, Gumina RJ, Liu SL. 2022. Evasion of neutralizing antibody responses by the SARS-CoV-2 BA.2.75 variant. Cell Host Microbe 30:1518–1526 e4.

31. Sheward DJ, Kim C, Fischbach J, Sato K, Muschiol S, Ehling RA, Bjorkstrom NK, Karlsson Hedestam GB, Reddy ST, Albert J, Peacock TP, Murrell B. 2022. Omicron sublineage BA.2.75.2 exhibits extensive escape from neutralising antibodies. Lancet Infect Dis 22:1538–1540.

32. Amato L, Jurisic L, Puglia I, Di Lollo V, Curini V, Torzi G, Di Girolamo A, Mangone I, Mancinelli A, Decaro N, Calistri P, Di Giallonardo F, Lorusso A, D’Alterio N. 2021. Multiple detection and spread of novel strains of the SARS-CoV-2 B.1.177 (B.1.177.75) lineage that test negative by a commercially available nucleocapsid gene real-time RT-PCR. Emerg Microbes Infect 10:1148–1155.

33. Holland SC, Bains A, Holland LA, Smith MF, Sullins RA, Mellor NJ, Thomas AW, Johnson N, Murugan V, Lim ES. 2022. SARS-CoV-2 Delta Variant N Gene Mutations Reduce Sensitivity to the TaqPath COVID-19 Multiplex Molecular Diagnostic Assay. Viruses 14.

34. Vanaerschot M, Mann SA, Webber JT, Kamm J, Bell SM, Bell J, Hong SN, Nguyen MP, Chan LY, Bhatt KD, Tan M, Detweiler AM, Espinosa A, Wu W, Batson J, Dynerman D, Wadford DA, Puschnik AS, Neff N, Ahyong V, Miller S, Ayscue P, Tato CM, Paul S, Kistler AL, DeRisi JL, Crawford ED. 2020. Identification of a Polymorphism in the N Gene of SARS-CoV-2 That Adversely Impacts Detection by Reverse Transcription-PCR. J Clin Microbiol 59.

35. National Institutes of Health. 2022. Anti-SARS-CoV-2 Monoclonal Antibodies. https://www.covid19treatmentguidelines.nih.gov/therapies/antivirals-including-antibody-products/anti-sars-cov-2-monoclonal-antibodies/. Accessed

36. US FDA. 2022. Coronavirus (COVID-19) Update: FDA Limits Use of Certain Monoclonal Antibodies to Treat COVID-19 Due to the Omicron Variant. https://www.fda.gov/news-events/press-announcements/coronavirus-covid-19-update-fda-limits-use-certain-monoclonal-antibodies-treat-covid-19-due-omicron. Accessed 2022-12-10.

37. Heilmann E, Costacurta F, Volland A, von Laer D. 2022. SARS-CoV-2 3CLpro mutations confer resistance to Paxlovid (nirmatrelvir/ritonavir) in a VSV-based, non-gain-of-function system. BioRxiv doi:10.1101/2022.07.02.495455.

38. Rockett R, Basile K, Maddocks S, Fong W, Agius JE, Johnson-Mackinnon J, Arnott A, Chandra S, Gall M, Draper J, Martinez E, Sim EM, Lee C, Ngo C, Ramsperger M, Ginn AN, Wang Q, Fennell M, Ko D, Lim HL, Gilroy N, O’Sullivan Mvn, Chen SC, Kok J, Dwyer DE, Sintchenko V. 2022. Resistance Mutations in SARS-CoV-2 Delta Variant after Sotrovimab Use. N Engl J Med 386:1477–1479.

39. Hwang HS, Lo CM, Murphy M, Grudda T, Gallagher N, Luo CH, Robinson ML, Mirza A, Conte M, Conte A, Zhou R, Vergara C, Brooke CB, Pekosz A, Mostafa HH, Manabe YC, Thio CL, Balagopal A. 2022. Characterizing SARS-CoV-2 transcription of subgenomic and genomic RNAs during early human infection using multiplexed droplet digital PCR. J Infect Dis doi:10.1093/infdis/jiac472.

