## Supplementary materials for "Digital PCR discriminates between SARS-CoV-2 Omicron variants and immune escape mutations"

Supplemental material

Table S1: Digital PCR probe sequences and modifications

| Probe | Sequence | 5’ Fluorescent dye | 3’ modification |
|---|---|---|---|
| Orf1ab3395_BA1_FAM | CTATGAGGCACAATT | FAM | MGBNFQ |
| Orf1ab3395_Delta_VIC | CTATGAGGCCCAATT | VIC | MGBNFQ |
| Orf1ab3395_BA2_NED | CTATGAGACACAATT | NED | MGBNFQ |
| S143_BA.1_FAM | CATTTTTGGACCACAAAAA | FAM | MGBNFQ |
| S143_DeltaBA2_VIC | GGATGTTTATTACCACA | VIC | MGBNFQ |
| R346_JUN | ACGCCACCAGATTTGCA | JUN | QSY |
| R346T_ABY | ACGCCACCACATTTGCA | ABY | QSY |
| K444_VIC | GCTTGATTCTAAGGTT | VIC | MGBNFQ |
| K444T_FAM | GCTTGATTCTACGGTT | FAM | MGBNFQ |
| N460_FAM | GAAGTCTAATCTCAACC | FAM | MGBNFQ |
| N460Kaag_VIC | GAAGTCTAAACTCAAAC | VIC | MGBNFQ |
| N460Kaaa_VIC | GAAGTCTAAGCTCAAAC | VIC | MGBNFQ |
| F486_ABY | TGTTGCAGGTTTTAATTGTTACT | ABY | QSY |
| F486S_JUN | TGTTGCAGGTTCTAATTGTTACT | JUN | QSY |
| F486V_JUN | TGTTGCAGGTGTTAATTGTTACT | JUN | QSY |

Table S2: Digital PCR primer sequences

| Primer | Sequence |
|---|---|
| Orf1ab3395_fwd | TGTTAGCTTGTTACAATGGTTCACC |
| Orf1ab3395_rev | CCACATGAACCATTAAGGAATGAACC |
| S143_fwd | GCTACTAATGTTGTTATTAAAGTCTGTGAATTTC |
| S143_rev | GTGCAATTATTCGCACTAGAATAAACTC |
| R346_fwd | CTAATATTACAAACTTGTGCCC |
| R346_rev | GGACAGAATAATCAGCAACA |
| K444_N460_fwd | GCTGCGTTATAGCTTGG |
| K444_N460_rev | ATAGATTTCAGTTGAAATATCTCTC |
| F486_fwd | GGCCGGTAACAAACC |
| F486_rev | TACTACTACTCTGTATGGTTGG |

Table S3: Digital PCR synthetic DNA templates

| Name | Sequence | Mutations from Wuhan1 (probe-specific mutations in bold) |
|---|---|---|
| Orf1ab3395_Delta | CGCTGTAATACGACTCACTATAGGGTTAAGGT<br>TGATACAGCCAATCCTAAGACACCTAAGTATAA<br>GTTTGTTCGCATTCAACCAGGACAGACTTTTTC<br>AGTGTTAGCTTGTTACAATGGTTCACCATCTG<br>GTGTTTACCAATGTGCTATGAGGCCCAATTTC<br>ACTATTAAGGGTTCATTCCTTAATGGTTCATGT<br>GGTAGTGTTGGTTTTAACATAGATTATGACTGT<br>GTCTCTTTTTGTTACATGCACCATATGGAATTA<br>CCAACTGGAGTTCATGCTGGCACAGACTTA | None |
| Orf1ab3395_BA1 | CGCTGTAATACGACTCACTATAGGGTTAAGGT<br>TGATACAGCCAATCCTAAGACACCTAAGTATAA<br>GTTTGTTCGCATTCAACCAGGACAGACTTTTTC<br>AGTGTTAGCTTGTTACAATGGTTCACCATCTG<br>GTGTTTACCAATGTGCTATGAGGCACAATTTCA<br>CTATTAAGGGTTCATTCCTTAATGGTTCATGTG<br>GTAGTGTTGGTTTTAACATAGATTATGACTGTG<br>TCTcTTTTTGTTACATGCACCATATGGAATTAC<br>CAACTGGAGTTCATGCTGGCACAGACTTA | **P3395H** |
| Orf1ab3395_BA2 | CGCTGTAATACGACTCACTATAGGGTTAAGGTT<br>GATACAGCCAATCCTAAGACACCTAAGTATAAGTTTGTT<br>CGCATTCAACCAGGACAGACTTTTTCAGTGTTAGCTTGT<br>TACAATGGTTCACCATCTGGTGTTTACCAATGTGCTATG<br>AGACACAATTTCACTATTAAGGGTTCATTCCTTAATGGT<br>TCATGTGGTAGTGTTGGTTTTAACATAGATTATGACTGT<br>GTCTCTTTTTGTTACATGCACCATATGGAATTACCAACT<br>GGAGTTCATGCTGGCACAGACTTA | **a10447g** (Synonymous R3394), **P3395H** |
| S143_Delta/BA.2 | CGCTGTAATACGACTCACTATAGGGAGAGGCTG<br>GATTTTTGGTACTACTTTAGATTCGAAGACCCAGTCCCT<br>ACTTATTGTTAATAACGCTACTAATGTTGTTATTAAAGT<br>CTGTGAATTTCAATTTTGTAATGATCCATTTTTGGATGTT<br>TATTACCACAAAAACAACAAAAGTTGGATGGAAAGTG<br>GAGTTTATTCTAGTGCGAATAATTGCACTTTTGAATATG<br>TCTCTCAGCCTTTTCTTATGGACCTTGAAGGAAAACAGG<br>GTAATTTCAAAAA | |
| S143_BA1 | CGCTGTAATACGACTCACTATAGGGTAATAAGAGGCTG<br>GATTTTTGGTACTACTTTAGATTCGAAGACCCAGTCCCT<br>ACTTATTGTTAATAACGCTACTAATGTTGTTATTAAAGT<br>CTGTGAATTTCAATTTTGTAATGATCCATTTTTGGACCA<br>CAAAAACAACAAAAGTTGGATGGAAAGTGAGTTCAGA<br>GTTTATTCTAGTGCGAATAATTGCACTTTTGAATATGTC | **G142D**, **del143-145** |

| | | |
|---|---|---|
| | TCTCAGCCTTTTCTTATGGACCTTGAAGGAAAACAGGG<br>TAATTTCAAAAATCTT | |
| RBD_Ref-like | TAATACGACTCACTATAGGGAGATTAGATTTCCTAATAT<br>TACAAACTTGTGCCCTTTTGATGAAGTTTTTAACGCCAC<br>CAGATTTGCATCTGTTTATGCTTGGAACAGGAAGAGAA<br>TCAGCAACTGTGTTGCTGATTATTCTGTCCTATATAATTT<br>CGCACCATTTTTCGCTTTTAAGTGTTATGGAGTGTCTCC<br>TACTAAATTAAATGATCTCTGCTTTACTAATGTCTATGC<br>AGATTCATTTGTAATTAGAGGTAATGAAGTCAGCCAAA<br>TCGCTCCAGGGCAAACTGGAAATATTGCTGATTATAAT<br>TATAAATTACCAGATGATTTTACAGGCTGCGTTATAGCT<br>TGGAATTCTAACAAGCTTGATTCTAAGGTTGGTGGTAA<br>TTATAATTACCTGTATAGATTGTTTAGGAAGTCTAATCT<br>CAAACCTTTTGAGAGAGATATTTCAACTGAAATCTATCA<br>GGCCGGTAACAAACCTTGTAATGGTGTTGCAGGTTTTA<br>ATTGTTACTTTCCTTTACAATCATATGGTTTCCGACCCAC<br>TTATGGTGTTGGTCACCAACCATACAGAGTAGTAGTAC<br>TTTCTTTT | G339D, S371F, S373P, S375F, T376A, D405N, R408S, K417N, N440K, S477N, T478K, E484A, Q498R, N501Y, Y505H |
| RBD_Mutant1 | TAATACGACTCACTATAGGGAGATTAGATTTCCTAATAT<br>TACAAACTTGTGCCCTTTTGATGAAGTTTTTAACGCCAC<br>CACATTTGCATCTGTTTATGCTTGGAACAGGAAGAGAA<br>TCAGCAACTGTGTTGCTGATTATTCTGTCCTATATAATTT<br>CGCACCATTTTTCGCTTTTAAGTGTTATGGAGTGTCTCC<br>TACTAAATTAAATGATCTCTGCTTTACTAATGTCTATGC<br>AGATTCATTTGTAATTAGAGGTAATGAAGTCAGCCAAA<br>TCGCTCCAGGGCAAACTGGAAATATTGCTGATTATAAT<br>TATAAATTACCAGATGATTTTACAGGCTGCGTTATAGCT<br>TGGAATTCTAACAAGCTTGATTCTACGGTTGGTGGTAA<br>TTATAATTACCGGTATAGATTGTTTAGGAAGTCTAAACT<br>CAAACCTTTTGAGAGAGATATTTCAACTGAAATCTATCA<br>GGCCGGTAACAAACCTTGTAATGGTGTTGCAGGTTCTA<br>ATTGTTACTTTCCTTTACAATCATATGGTTTCCGACCCAC<br>TTATGGTGTTGGTCACCAACCATACAGAGTAGTAGTAC<br>TTTCTTTT | G339D, **R346T**, S371F, S373P, S375F, T376A, D405N, R408S, K417N, N440K, **K444T**, L452R, **N460K** (aaa codon), S477N, T478K, E484A, **F486S**, Q498R, N501Y, Y505H |
| RBD_Mutant2 | TAATACGACTCACTATAGGGAGATTAGATTTCCTAATAT<br>TACAAACTTGTGCCCTTTTGATGAAGTTTTTAACGCCAC<br>CACATTTGCATCTGTTTATGCTTGGAACAGGAAGAGAA<br>TCAGCAACTGTGTTGCTGATTATTCTGTCCTATATAATTT<br>CGCACCATTTTTCGCTTTTAAGTGTTATGGAGTGTCTCC<br>TACTAAATTAAATGATCTCTGCTTTACTAATGTCTATGC<br>AGATTCATTTGTAATTAGAGGTAATGAAGTCAGCCAAA<br>TCGCTCCAGGGCAAACTGGAAATATTGCTGATTATAAT<br>TATAAATTACCAGATGATTTTACAGGCTGCGTTATAGCT<br>TGGAATTCTAACAAGCTTGATTCTATGGTTGGTGGTAA<br>TTATAATTACCGGTATAGATTGTTTAGGAAGTCTAAGCT<br>CAAACCTTTTGAGAGAGATATTTCAACTGAAATCTATCA<br>GGCCGGTAACAAACCTTGTAATGGTGTTGCAGGTGTTA | G339D, **R346T**, S371F, S373P, S375F, T376A, D405N, R408S, K417N, N440K, K444M, L452R, **N460K** (aag codon), S477N, T478K, E484A, **F486V**, Q498R, N501Y, Y505H |

ATTGTTACTTTCCTTTACAATCATATGGTTTCCGACCCAC
TTATGGTGTTGGTCACCAACCATACAGAGTAGTAGTAC
TTTCTTTT

### Supplemental Figure S1

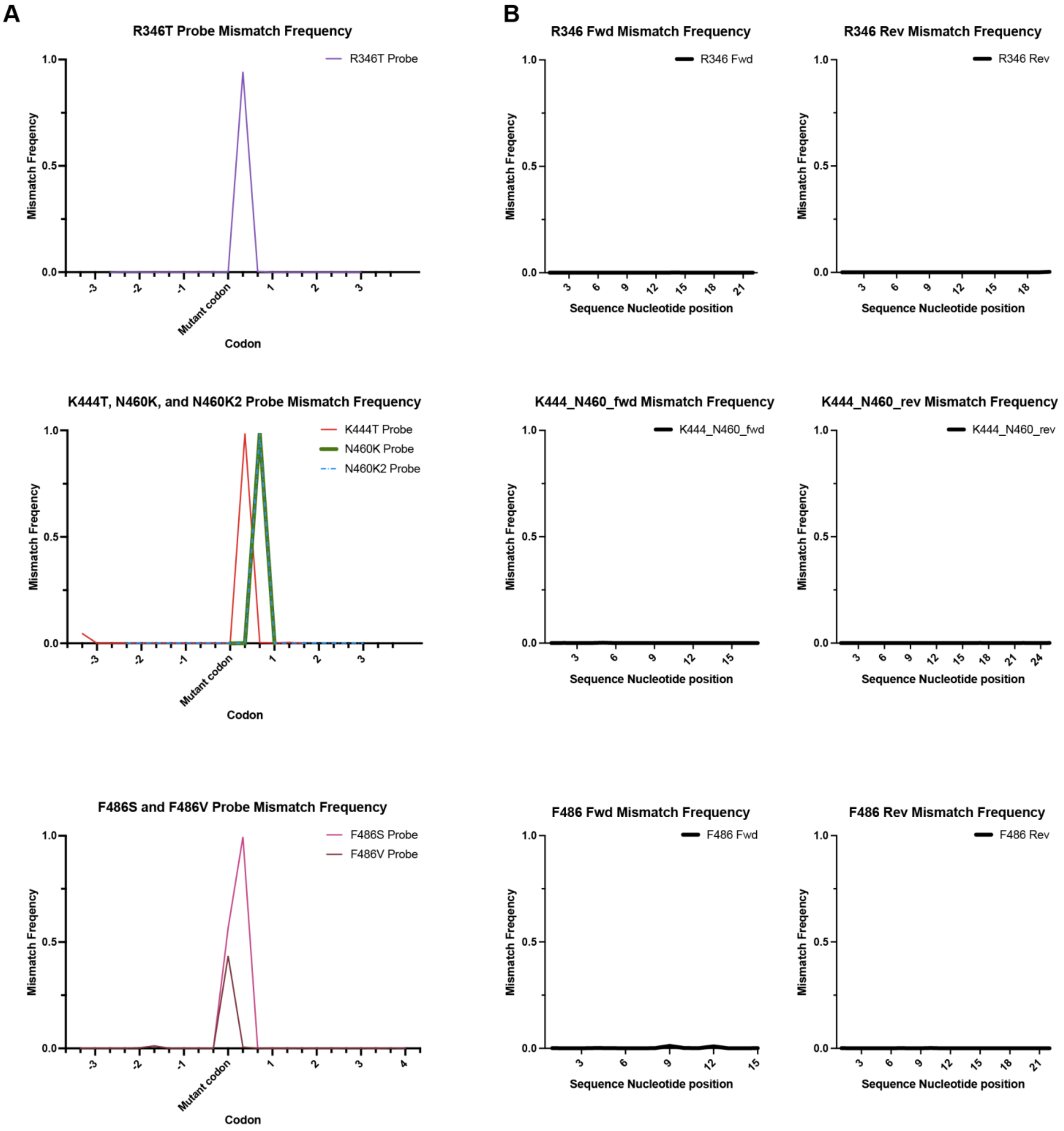


Fig S1: Failure frequency of probes (A) and primers (B) used in the *spike* RBD digital PCR assays.
